# Analyzing the Global Impact of COVID-19 Vaccination Progress: A Result-oriented Storytelling Approach

**DOI:** 10.1101/2021.03.26.21254432

**Authors:** Samrat Kumar Dey, Md. Mahbubur Rahman, Umme Raihan Siddiqi, Arpita Howlader, Arifuzzaman Tushar

## Abstract

The next big step in combating the coronavirus disease 2019 (COVID-19) pandemic will be gaining widespread acceptance of a vaccination campaign for severe acute respiratory syndrome coronavirus 2 (SARS-CoV-2), but achieving high uptake need proper understandings. Many health professionals, researchers, statisticians, and programmers to track the viruses spread in different parts of the world have used various methods. However, the proliferation of vaccines produced by talented scientists around the world has sparked a strong desire to extract meaningful insights from available data. Until now, several vaccines against coronavirus disease (COVID-19) have been approved and are being distributed worldwide in various regions. This study aims to report the detailed data analysis and result-oriented storytelling of the COVID-19 vaccination program of different countries across the globe. To analyze the vaccination trend globally this research utilized two different open datasets provided by ourworldindata.org and worldometers.info. An exploratory data analysis (EDA) with interactive data visualization using various python libraries was conducted, and the results are presented in this article to better understand the impact of ongoing vaccination programs around the world. Apart from the valuable insights gained from the data of various countries, this investigation also included a comparison of the number of confirmed and death cases before and after vaccination to determine the efficacy of each vaccine in each country. The results show that a large number of people are still undecided about whether or not to get a COVID-19 vaccine, despite the virus’s continued devastating effects on communities. Overall, our findings contribute to ongoing research aimed at informing policy on how to persuade the unvaccinated to be vaccinated.

## 1. Introduction

Over the history of humankind, vaccine played a crucial role to overcome epidemic situations. Starting with deliberate variolation back in the 10^th^ century to our modern third-generation (RNA vaccines and DNA vaccines) **[1] [2]** vaccination helped us to fight back against deadly viruses and sustain the human race. Since SARS-CoV-2 is a highly contagious virus that affects populations all over the world, vaccines are the most important public health measure and the most effective strategy for protecting the population from COVID-19. Beginning in a fish market in Wuhan, Hubei Province, China, in December 2019, a novel coronavirus (SARS-CoV-2) spread around the world, causing COVID-19 disease in millions of people **[3]**. The World Health Organization (WHO) declared COVID-19 on March 11, 2020, for the sixth time in history, due to its high human-to-human transmission rate **[4]**. According to the statistics of worldometer.info, more than 120 million people had been infected as of March 15, 2021, with 2.66 million deaths and 20.79 million active cases. The race to develop COVID19 vaccines to combat the disease’s spread and disastrous consequences is still on, and new, more effective vaccines are likely to emerge as the pandemic progresses **[5] [6]**. Several vaccines to prevent COVID-19 infection were approved in December 2020, and more than 50 COVID-19 vaccine candidates were being produced **[7] [8]**. The WHO has identified 48 vaccine candidates that are currently being tested in clinical trials **[9]**. The first step in developing a vaccine for any virus is to determine its genetic sequence. However, previously, on December 31, 2019, the World Health Organization issued a warning about a new coronavirus strain that is affecting people in China **[10]**. Later, officials from the China confirmed the identification of a new type of coronavirus infecting human bodies and the first genetic sequence for SARS-CoV-2 was released on January 11, 2020. Non-pharmaceutical interventions are used to minimize transmission and the burden of coronavirus disease 2019 (COVID-19) in the absence of safe and highly efficient vaccines and treatment options, but most of these interventions have high economic costs **[11]**. To reduce the substantial burden of COVID-19 morbidity and mortality, effective COVID-19 vaccines are desperately needed. Vaccine development is a lengthy process that necessitates numerous testing phases to ensure adequate safety and immunogenicity in a variety of people (i.e., different ages, medical conditions, severity of attack, geographic location etc.). According to the National Center for Biotechnology Information (NCBI), a vaccine must go through four stages of clinical trials before receiving a license to produce it, which can take up to a decade. However, due to the urgent pandemic situation around the world, the COVID-19 vaccine development process was shortened to 12-18 months while retaining safety and effectiveness standards. A number of factors, including a monumental breakthrough in biotechnology and molecular biology, as well as a collaboration between government and private research institutes, enables the extreme compression of the COVID-19 vaccine development process. The pandemic’s humanitarian and economic consequences are guiding the growth of next-generation vaccine technology platforms around the world. As a result of the COVID-19 vaccine’s production being accelerated, the first candidate entered human clinical trials with unparalleled speed on March 16, 2020 **[12]**. As of March 23, 2021, there are 349 COVID-19 treatment (drug medicine) methods in trials, and 267 vaccines are in progress, with 83 in various stages of clinical testing **[9] [13]**. Until 25 March 2021 150 countries from a different region of the world using vaccine from 11 different manufacturers (4 authorized and 7 with emergency use permission) to vaccinate their citizens. **Table 1** summarizes the various stages of vaccine development process.

**Table 1:**
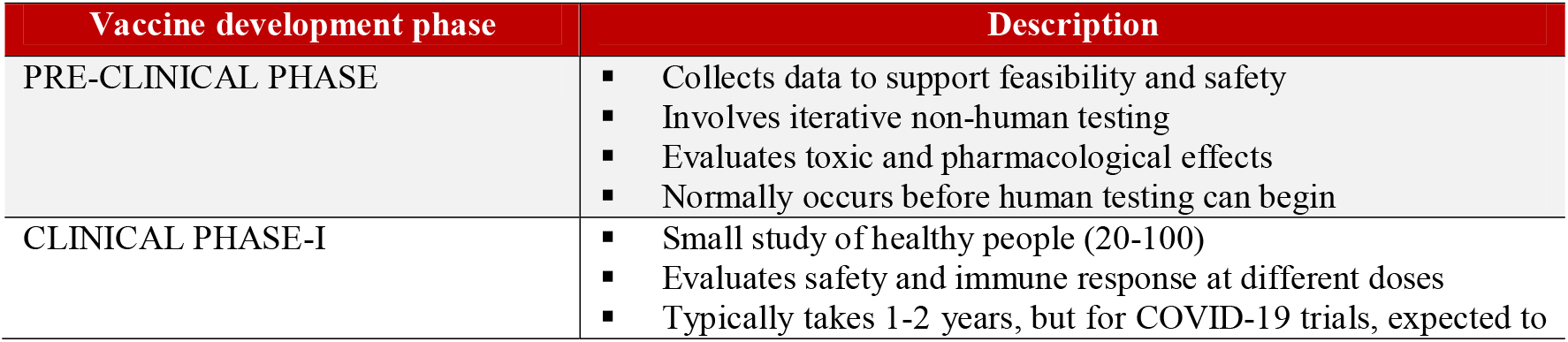
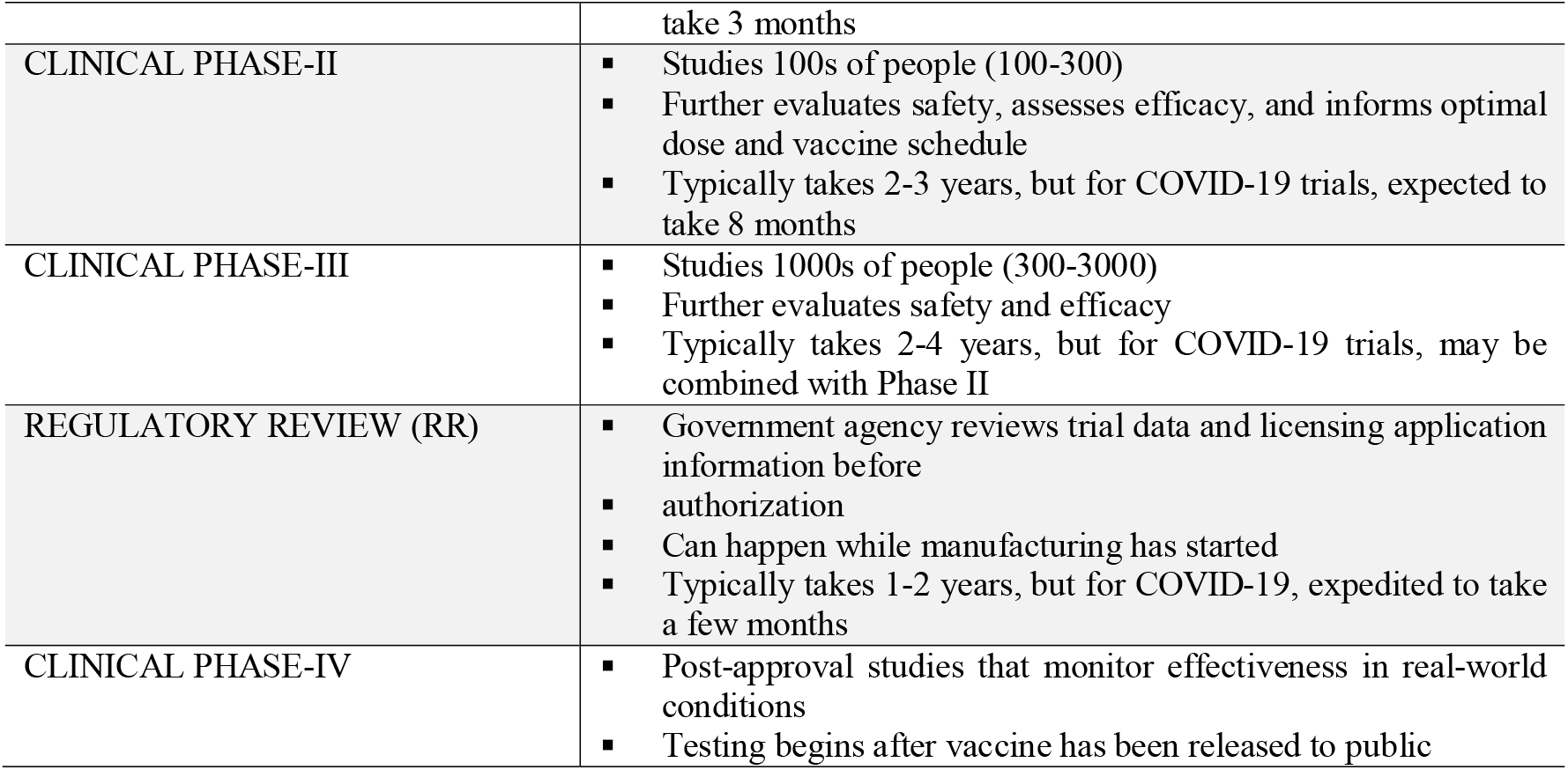
Phases of vaccine development and descriptions of each **[14]**

| Vaccine development phase | Description |
| --- | --- |
| PRE-CLINICAL PHASE | Collects data to support feasibility and safety<br>Involves iterative non-human testing<br>Evaluates toxic and pharmacological effects<br>Normally occurs before human testing can begin |
| CLINICAL PHASE-I | Small study of healthy people (20-100)<br>Evaluates safety and immune response at different doses<br>Typically takes 1-2 years, but for COVID-19 trials, expected to |
|  | take 3 months |
| CLINICAL PHASE-II | Studies 100s of people (100-300)<br>Further evaluates safety, assesses efficacy, and informs optimal dose and vaccine schedule<br>Typically takes 2-3 years, but for COVID-19 trials, expected to take 8 months |
| CLINICAL PHASE-III | Studies 1000s of people (300-3000)<br>Further evaluates safety and efficacy<br>Typically takes 2-4 years, but for COVID-19 trials, may be combined with Phase II |
| REGULATORY REVIEW (RR) | Government agency reviews trial data and licensing application information before<br>authorization<br>Can happen while manufacturing has started<br>Typically takes 1-2 years, but for COVID-19, expedited to take a few months |
| CLINICAL PHASE-IV | Post-approval studies that monitor effectiveness in real-world conditions<br>Testing begins after vaccine has been released to public |

To the best of our knowledge, no previously published work has focused on the progress on COVID-19 vaccination worldwide with the support of interactive Exploratory Data Analysis (EDA) and visualizations. The present study aim to evaluate the global participation of citizens by analyzing and highlighting different characteristics towards successful vaccination. The primary objectives of this exploration is to answers different query regarding the progress of vaccination to understand the ongoing safe vaccination program around the globe. People’s experience, attitudes, and perceptions about COVID-19 vaccinations are critical for Government and policymakers to address all obstacles to vaccine distribution in this scenario. Furthermore, it will inspire and give courage to those people around the world who oppose taking the vaccine for themselves.

## 2. Materials and Methods

This section will discuss about the different materials that we employed to build our methods. In this research, we have used the COVID-19 World Vaccination Progress dataset **[15]** available at Kaggle that tracks the Daily and Total Vaccination for COVID-19 in the World. However, COVID-19 World Vaccination Progress datasets is collected, merged, and updated regularly from Our World in Data GitHub repository (https://github.com/owid/covid-19-data). Following **Table 2** highlights the contents (data type, column name, and data description) of COVID-19 World Vaccination Progress Dataset. The dataset contain 15 different column (9 decimal, 3 string, 1 country and 2 other types of data) to track the progress of global vaccination of COVID-19 around the world. This exploration also utilized another dataset (Population by Country - 2020), available at Kaggle inherited from the Worldometer [16]. This dataset contains the information of 235 countries along with their population and there are 11 columns each representing different features of countries.

**Table 2:** Dataset details to keep track of COVID-19 vaccination rates around the world on a daily and total basis.

| Data type | Data Column | Data Description |
| --- | --- | --- |
| Country | country | Vaccination data is available for this country. |
| Country ISO Code | iso_code | Specific ISO code for the country |
| Date | date | Data entry date |
| Total number of vaccinations | total_vaccinations | This is the total number of immunizations in the country in its entirety; |
| Total number of people vaccinated | people_vaccinated | Depending on the immunization plan, a person may receive one or more (usually two) vaccines; at any given time, the number of vaccines can exceed the number of people. |
| Total number of people fully vaccinated | people_fully_vaccinated | This is the number of people who received the whole package of immunizations according to the immunization program (typically 2); at any given time, there might be one group of people who received one vaccine and another group (smaller) who received all vaccines in the scheme. |
| Daily vaccinations (raw) | daily_vaccinations_raw | The number of vaccinations for that date/country, or a particular data entry |
| Daily vaccinations | daily_vaccinations | The number of vaccinations for that date/country, or a particular data entry |
| Total vaccinations per hundred | total_vaccinations_per_hundred | Up to date in the world, the ratio (in percent) between the number of people who have been vaccinated and the total population |
| Total number of people vaccinated per hundred | people_vaccinated_per_hundred | Percentage of the population that has been immunized compared to the total population in the country |
| Total number of people fully vaccinated per hundred | people_fully_vaccinated_per_hundred | Ratio (in percent) of completely immunized population to total population up to date in the country |
| Daily vaccinations per million | daily_vaccinations_per_million | Proportion of completely immunized population to total population in the country (in percent) |
| Vaccines used in the country | vaccines | Vaccines used in the world as a whole (up to date) |
| Data source Authority | source_name | Source of the information (national authority, international organization, local organization etc.) |
| Data source website | source_website | website of the source of information |

### 2.1 Data Preparation and Cleaning

One of the important property of EDA before finding the insight form the data is to clean and prepare the data according to the requirements. Therefore, data cleaning is the most important step towards an effective data analysis. In our case, the dataset contains few “NaN” (not a number) values, some empty rows (having value 0), along with some redundant columns. By using and configuring the function of *df*.*drop* from pandas library, we changed “NaN” values to 0 and removed the entire row as per our requirements.

### 2.2 Data Visualization and Analysis

We analyzed our datasets with different Exploratory Data Analysis (EDA) **[17]** methods and visualize those outcome to provide analysis of different ongoing vaccination programs around the globe. For data ingestion, visualization and analysis purpose we initialized different python packages including NumPy (https://numpy.org/), Pandas (https://pandas.pydata.org/), Matplotlib (https://matplotlib.org/), Seaborn (https://seaborn.pydata.org/), and Plotly (https://plotly.com/). Mostly, we have used Seaborn for data visualization.

## 3. Results

This study analyzed the COVID-19 World Vaccination Progress dataset to convey the analysis of different ongoing vaccination programs around the globe. In this section the data analysis results and visualization will be described to find the answers to different query of this research including a) Finding out those countries who started vaccinating their citizens fastest, b) Those countries who have vaccinated the highest people, c) Different categories of offered vaccines, d) Name of the vaccines used by various countries, e) Finding daily vaccination trend of most vaccine used countries, and f) Vaccine impact analysis on new confirmed and death cases. To achieve the objectives of this research we have followed variety of data analysis and visualizations approach to highlight the results. Following **Figure 1**, represent the top 25 countries in the world who have started fastest vaccination in their country. Based on the bar chart, Israel leading the trend with more than 85 vaccine per hundred peoples and after that UAE ensured almost 70 vaccine for their peoples in per hundred. On other hand, England and UK almost ensure the same number of vaccine for their peoples.

**Figure 1:**
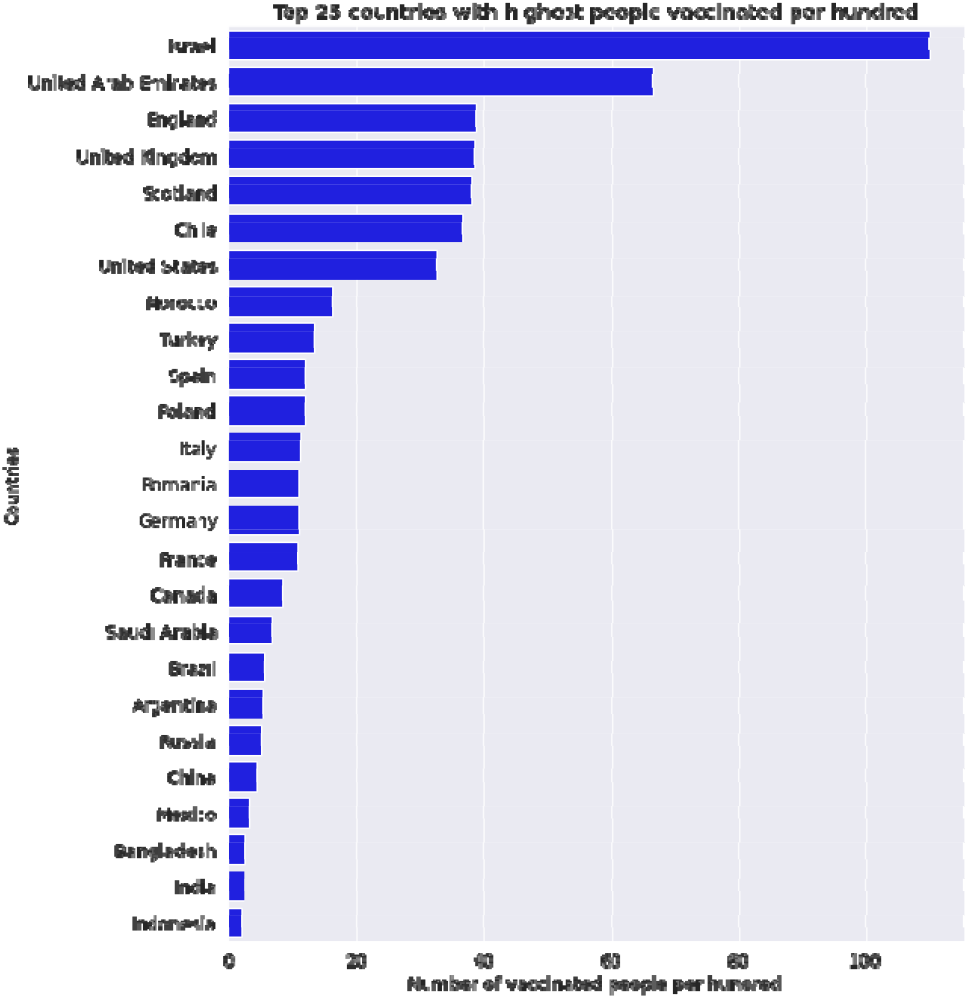
With more than 85 doses given for every 100 people, Israel clearly leads the world in terms of number of doses per head of population, as seen in the graph as of 17 March 2021.

Until 17 march, 2021 different scenario of worldwide vaccination progress worldwide has visualized in the **Figure 2**. Individual countrywide total vaccination, number of people who took vaccination for at least once, people who completed their double dose of vaccination and based on the population percentage of countrywide vaccination highlighted here. However, we have highlighted only top 25 countries for the better understandings of the entire scenario for each case.

**Figure 2:**
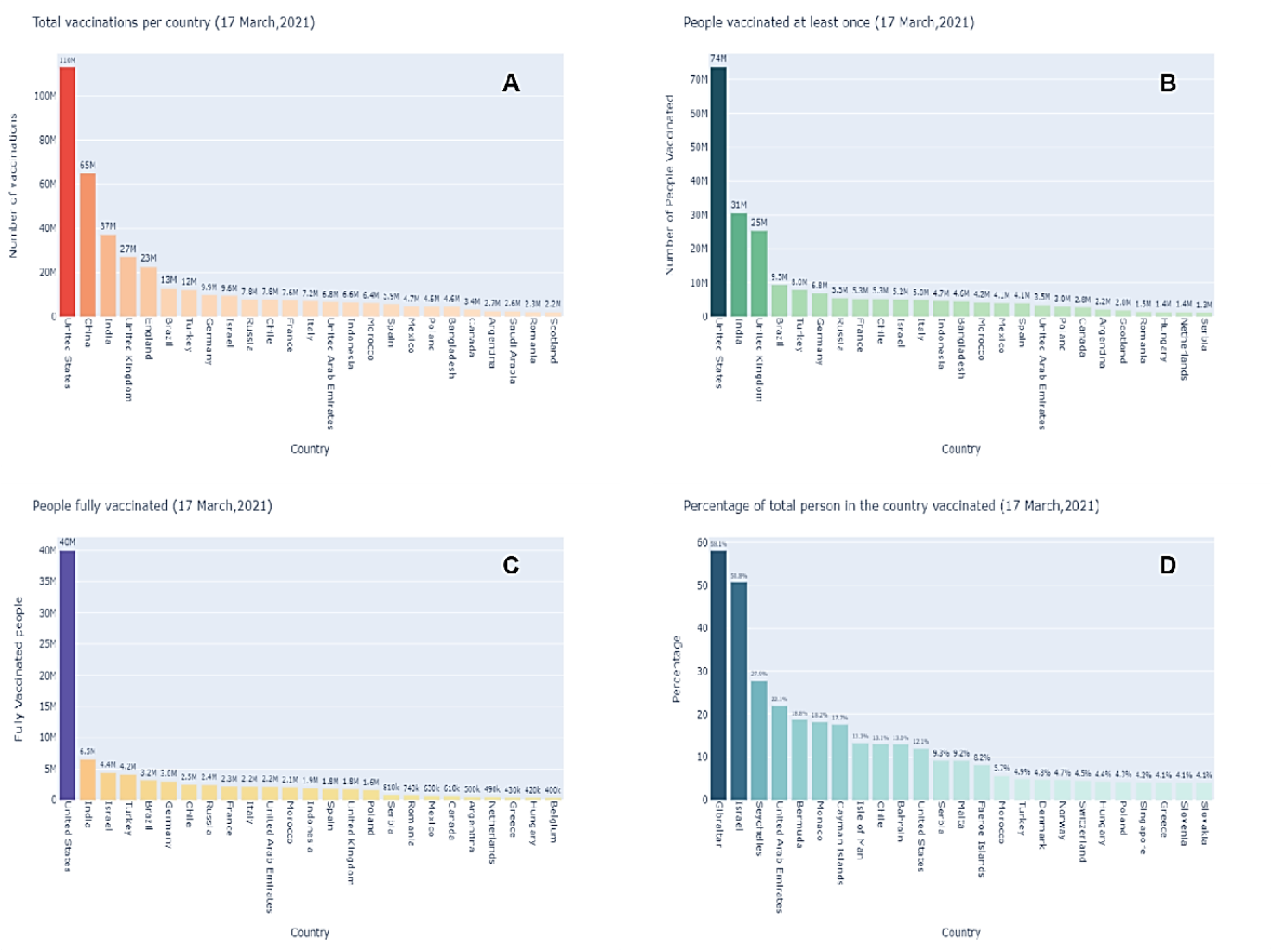
The bar plot visualization depicts different scenario of vaccination progress. 2(A) indicates that USA ensured110M vaccinations for their peoples and China after that with 65M until 17 March 2021. Those countries were individuals who have taken the vaccine for at least once is clearly depicted in 2(B). Surprisingly peoples of south Asian country like India already taken 31M vaccine and placing them in the second position after the USA. Successful countries those have managed to provide a complete dose (2 dose for each) for their people illustrates in 2(C). With 40M complete doses of vaccine, USA covers almost 12% of its total populations under fully vaccinated countries. Again, based on total population, 2(D) confirms that Gibraltar, Israel, and Seychelles vaccinated 58.11%, 50.78% and 27.87% of their peoples until 17 March 2021.

**Figure 3:**
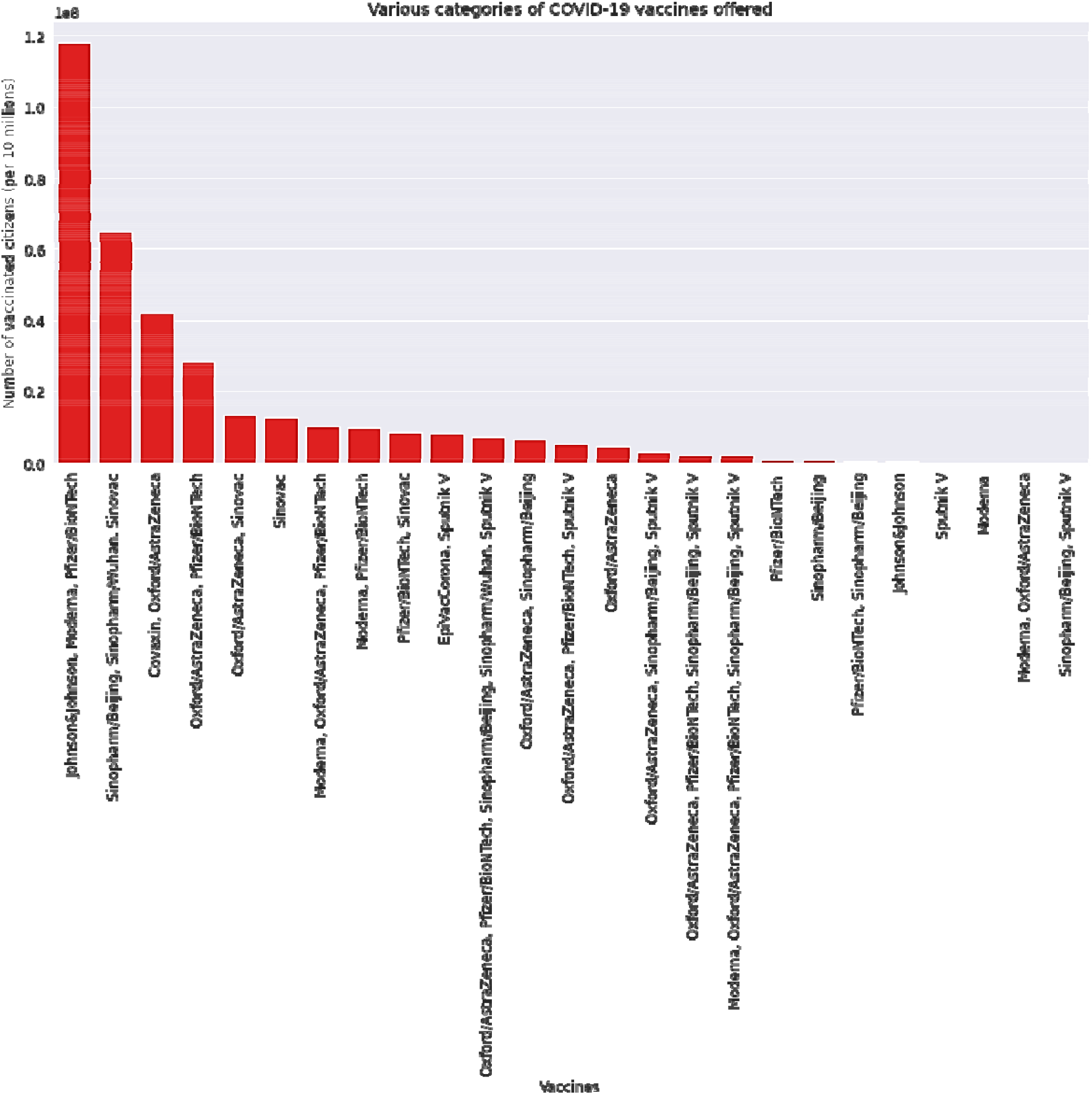
Pfizer tops the list of most-used COVID-19 vaccines in the world, followed by Mordema, and Oxford–AstraZeneca

One hundred and thirty seven (137) countries utilizing 25 different vaccine based on various region all over the globe. A Seaborn bar plot representation highlights all the available vaccine used in per 10 million until 17 March 2021 worldwide. Pfizer tops the list of most-used COVID-19 vaccines in the world, followed by Mordema, and Oxford–AstraZeneca. China mostly uses Sinovac vaccine and India uses Covaxin, Covishield for vaccinating its citizens.

A Plotly map representation of how different vaccines categories have been used by different countries around the globe is shown in **Figure 4**. United States mostly used Johnson&Johnson, Moderna, Pfizer/BioNTech vaccine whereas Canada ensured Moderna, Oxford/AstraZeneca, Pfizer/BioNTech in the North American Region. Furthermore, to provide an insight of Top 20 countries based on highest number of vaccine used, **Table 3** is designed based on the map to highlight the countries and their use of different vaccine for their citizens.

**Table 3:** Top 20 countries and the used of different vaccine for their citizen based on highest number of vaccine used for their citizens. Serial Number indicating the position of each countries on their usages of total number of vaccines.

| Sl. no. | Countries | Vaccine Used | Sl. no. | Countries | Vaccine Used |
| --- | --- | --- | --- | --- | --- |
| 1. | USA | Johnson&Johnson, Moderna, Pfizer/BioNTech | 2. | China | Sinopharm/Beijing, Sinopharm/Wuhan, Sinovac |
| 3. | India | Covaxin, Oxford/AstraZeneca | 4. | UK | Oxford/AstraZeneca, Pfizer/BioNTech |
| 5. | England | Oxford/AstraZeneca, Pfizer/BioNTech | 6. | Turkey | Sinovac |
| 7. | Brazil | Oxford/AstraZeneca, Sinovac | 8. | Germany | Moderna, Oxford/AstraZeneca, Pfizer/BioNTech |
| 9. | Israel | Moderna, Pfizer/BioNTech | 10. | Russia | EpiVacCorona, Sputnik V |
| 11. | Chile | Pfizer/BioNTech, Sinovac | 12. | France | Moderna, Oxford/AstraZeneca, Pfizer/BioNTech |
| 13. | Italy | Moderna, Oxford/AstraZeneca, Pfizer/BioNTech | 14. | Morocco | Oxford/AstraZeneca, Sinopharm/Beijing |
| 15. | Indonesia | Sinovac | 16. | UAE | Oxford/AstraZeneca, Pfizer/BioNTech, Sinopharm/Beijing, Sinopharm/Wuhan, Sputnik V |
| 17. | Spain | Moderna, Oxford/AstraZeneca, Pfizer/BioNTech | 18. | Poland | Moderna, Oxford/AstraZeneca, Pfizer/BioNTech |
| 19. | Mexico | Oxford/AstraZeneca, Pfizer/BioNTech, Sputnik V | 20. | Bangladesh | Oxford/AstraZeneca |

**Figure 4:**
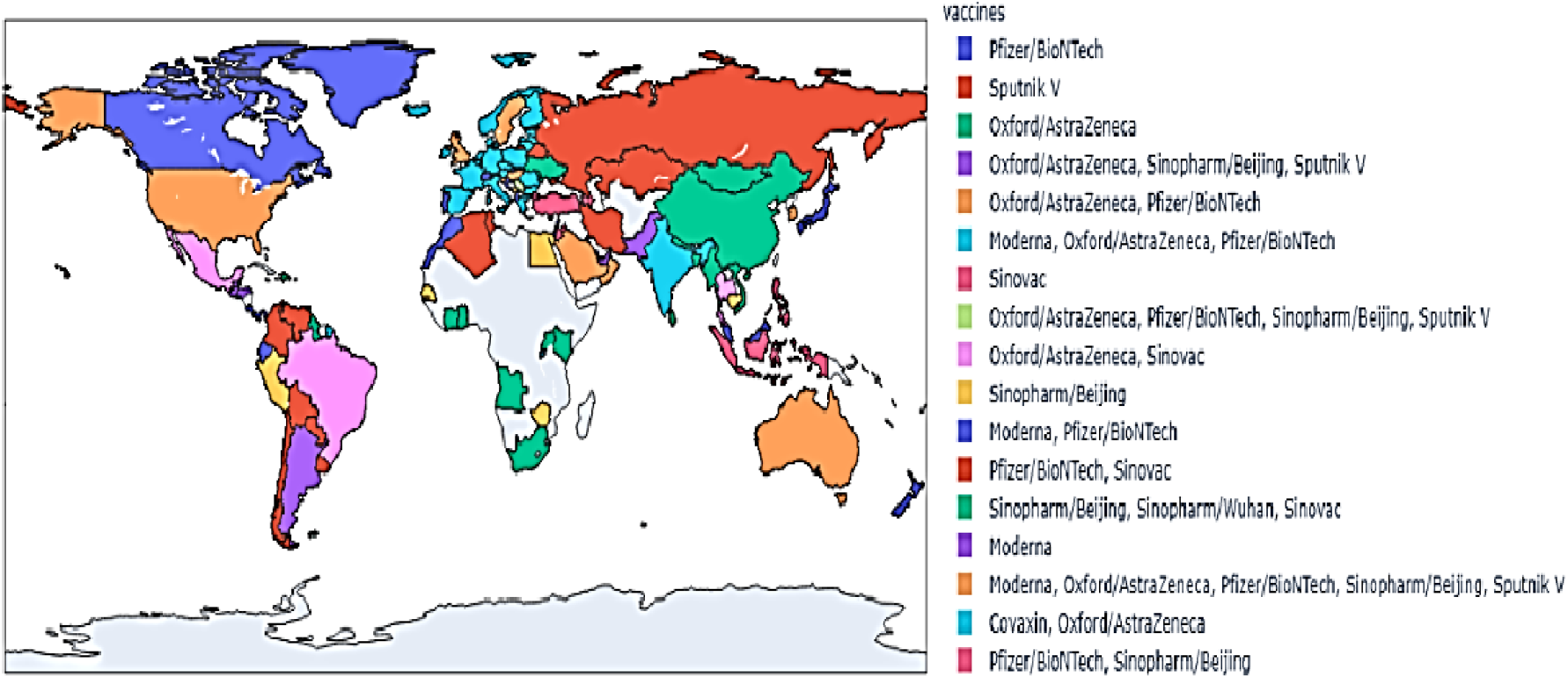
Map representation of different vaccines used by different countries viz., India□-□Covaxin, Oxford/AstraZeneca, USA□-□Moderna, Pfizer/BioNTech, Israel□-□Moderna, Pfizer/BioNTech, UK-Oxford/AstraZeneca, Pfizer/BioNTech.

To highlight the most and least vaccinated countries and vaccine used we further visualize that data in **Figure 5**. Graphics library of Plotly offers Sunburst plots (*px*.*sunburst*) that visualize the hierarchical data spanning outwards radially from root to leaves. Based on the ED analysis and visualization it confirms that United States mostly uses Johnson & Johnson, Moderna, and Pfizer/BioNTech vaccine with a rate of 32.62% (total vaccination= 109081860.0) in terms of total vaccinations until 17 March 2021. However, China and India are in next position respectively in the category of most vaccinated countries with a total vaccination of 64980000 (4.51%) and 32947432 (2.39%).

**Figure 5:**
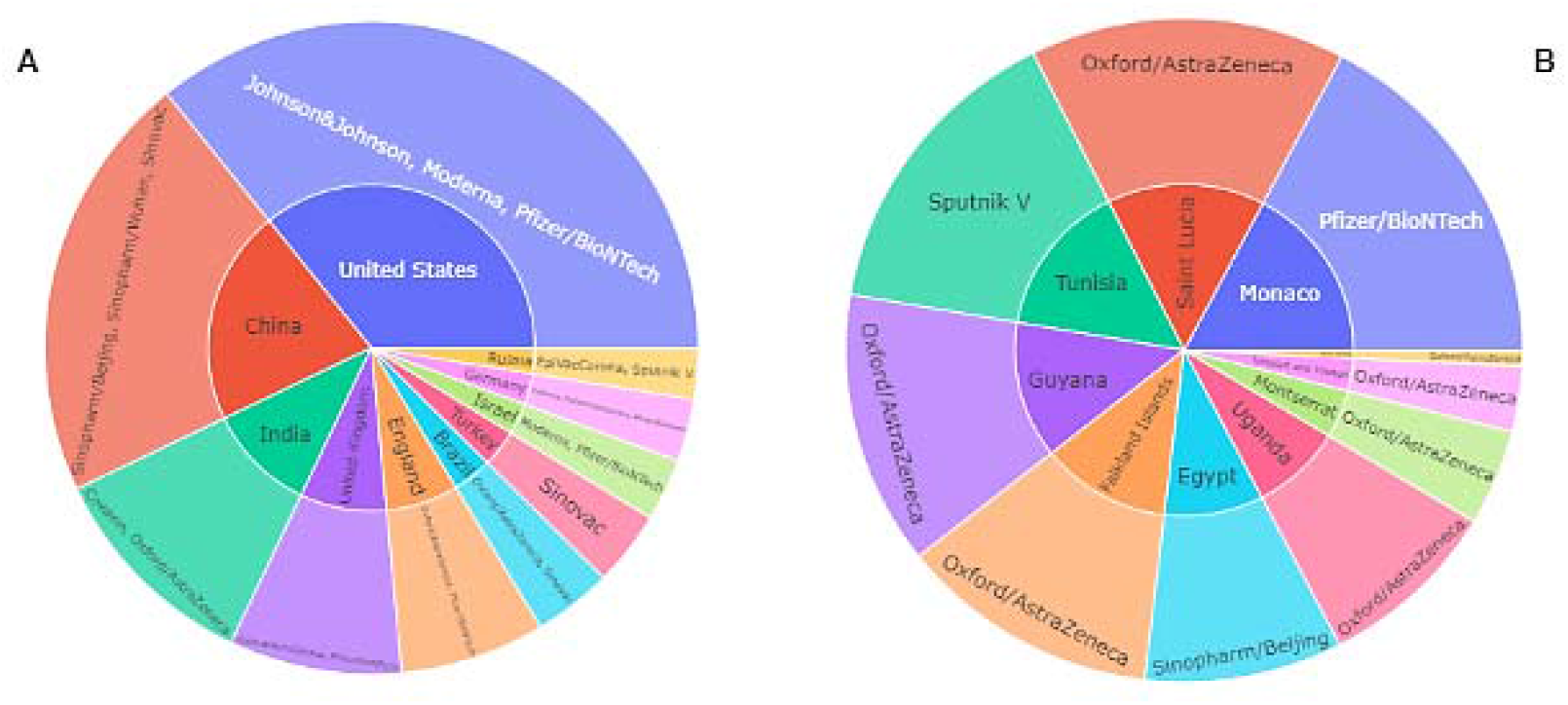
Sunburst plot representation of most and least vaccine used countries and their used vaccine. 5(A) shows the highest number of vaccine utilized countries along with the group of vaccine and 5(B) visualize the least vaccine used countries with their vaccine group utilizations.

However, for the countries that less vaccinated so far, Saint Helena only use the 107.0 vaccine of Oxford/AstraZeneca with total vaccination rate of 1.76%. Trinidad and Tobago and Montserrat remain in next position with a total vaccination of 440.0 (0.03%) and 652.0 (13.04%). Daily vaccinations is another important factor to assess the perception of people’s attitude towards successful vaccination. As a result, using data from the most vaccinated countries, we compiled a list of five (5) of the most vaccinated countries’ regular patterns from the start of vaccinations to the present **(Figure 6)**. United States, United Kingdom, China, India and England ensured highest number of vaccination in their country.

**Figure 6:**
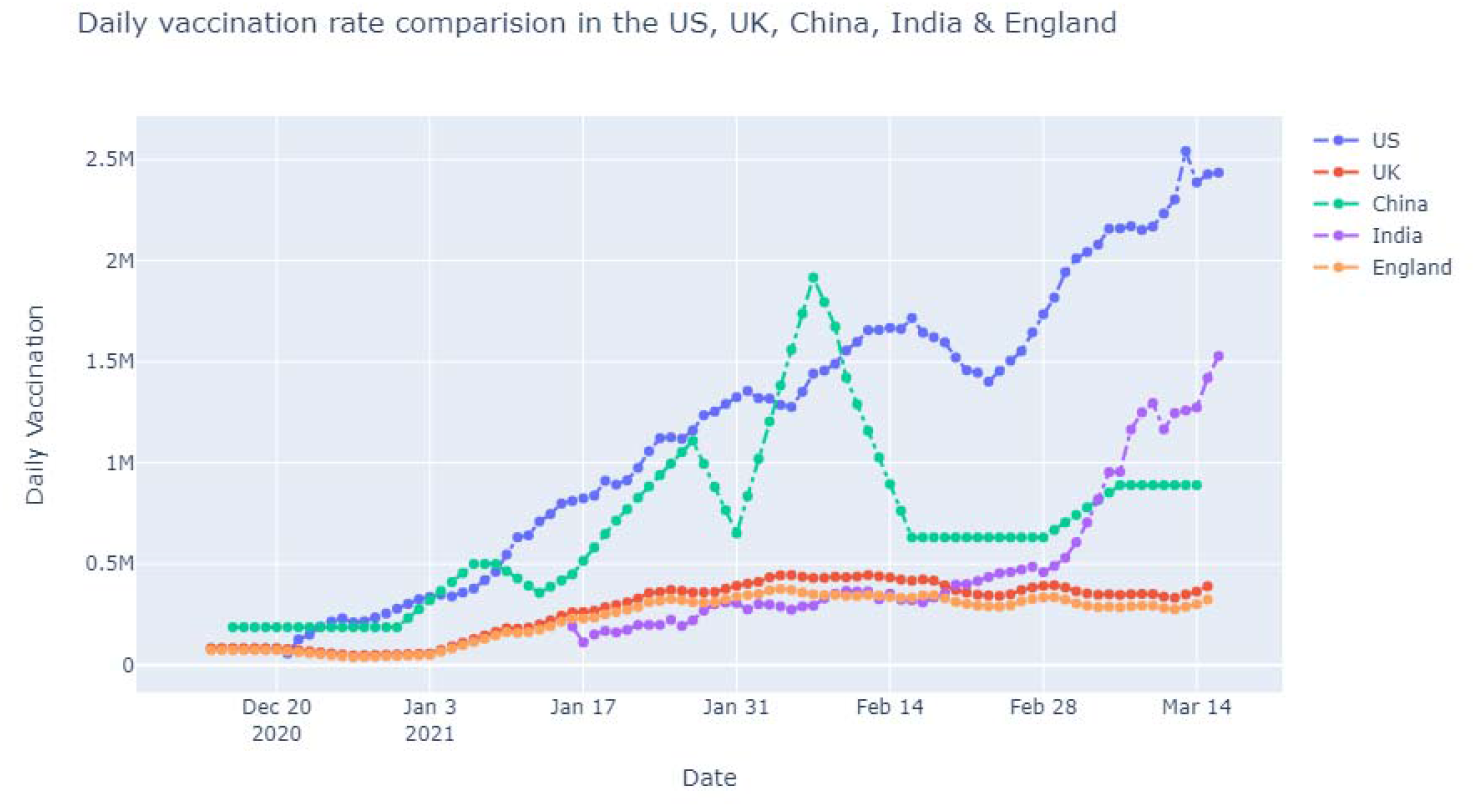
US, UK, China, India and England have vaccinated most people until now. Here, we have highlighted the daily vaccination trend for those countries. United States (US) leading the trend by vaccinating with almost 2.44 million peoples whereas England shows a downward curve by vaccinating 323.44K peoples exactly.

## 4. Discussion

This section will discuss on the findings of this exploration in more border way. **Table 4** contains details about vaccine categories and their current development stage. As of March 14, 2021, there are 251 vaccine in 10 different categories are in the process of development around the world whereas 11 vaccines including four (4) authorized and seven (7) from clinical phase-III are being used around the world **(Table 5)**.

**Table 4:** Variety of approaches among COVID-19 vaccine candidates until 14 March 2021

| Vaccine Category | Phase |  |  |  |  |  |  | Total |
| --- | --- | --- | --- | --- | --- | --- | --- | --- |
|  | Pre-Clinical | I | I/II | II | II/III | III | Authorized |  |
| DNA-based | 14 | 2 | 2 | - | 2 | 1 | - | 21 |
| Inactivated virus | 9 | 1 | 1 | 2 |  | 6 | - | 19 |
| Live attenuated virus | 3 | 1 | - | - | - | - | - | 4 |
| Non-replicating viral vector | 20 | 5 | 1 | - | - | 2 | 2 | 30 |
| Protein subunit | 62 | 7 | 7 | 3 | - | 3 | - | 82 |
| Replicating bacterial vector | 2 | - | - | - | - | - | - | 2 |
| Replicating viral vector | 18 | 2 | 2 | 1 | - | - | - | 23 |
| RNA-based vaccine | 25 | 2 | 2 | - | - | 1 | 2 | 32 |
| Unknown | 16 | - | - | - | - | - | - | 16 |
| Virus-like particle | 20 | - | 1 |  | 1 | - | - | 22 |
| **Total** | **189** | **20** | **16** | **6** | **3** | **13** | **4** | **251** |

**Table 5:**
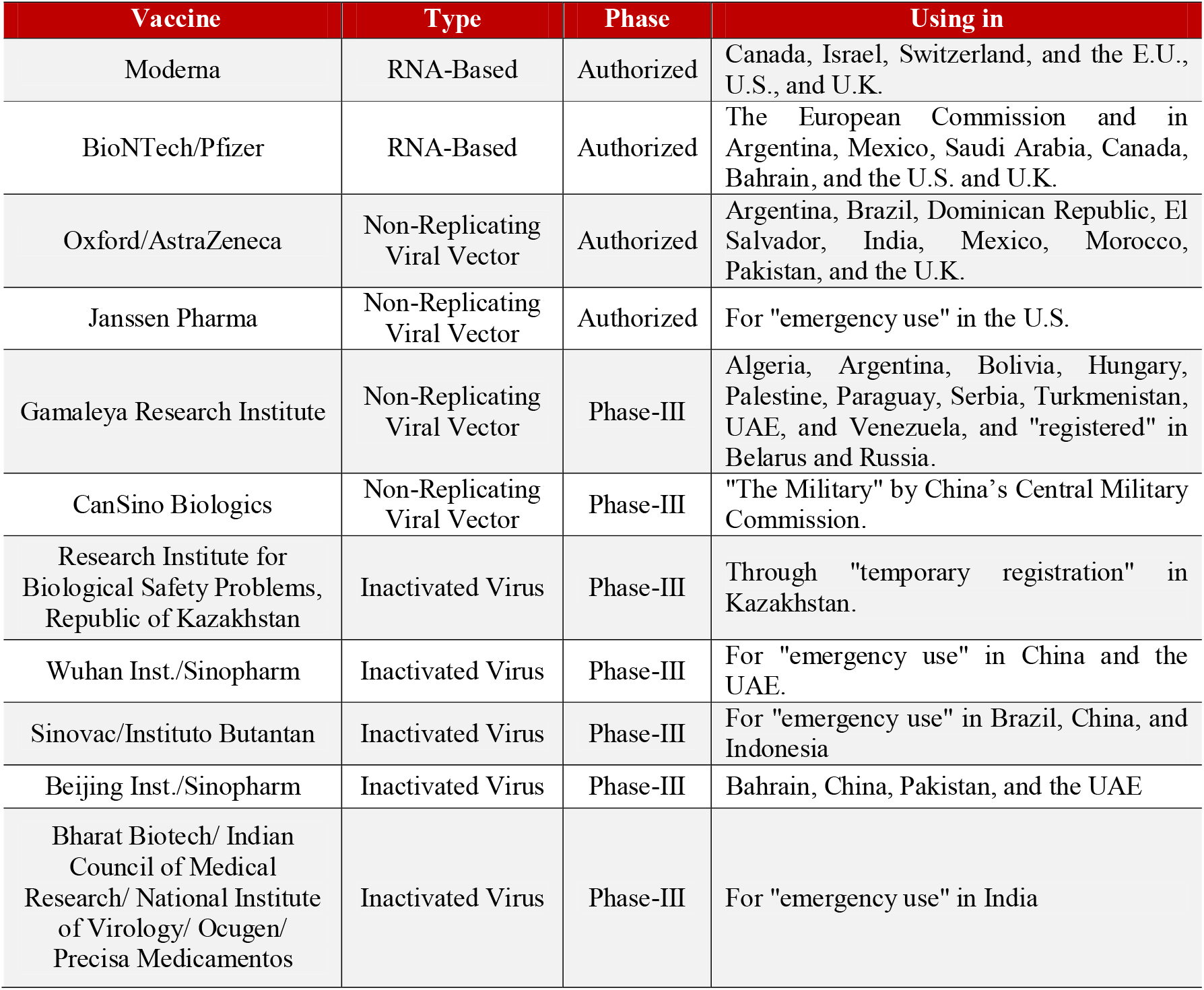
Wide categories of COVID-19 vaccines that are currently in use according to their type and phase.

Based on the results analysis and visualization many countries around the globe started applying vaccine to their citizens by the end of 2020. The rate of applying vaccines to the patients is highest in Isreal. It is because of its small size (in terms of both area and population), a relatively young population, relatively warm weather in December 2020, a centralized national system of government, and well-developed infrastructure for implementing prompt responses to large-scale national emergencies. Until 17 March 2021, The United States has the most vaccinated people of around 110M of its total population followed by China and India. As these are developed countries and they produce vaccine in their own lab; therefore, the accessibility of the vaccine is easier to its public. The impact of using vaccine on specific countries to reduce the spread of COVID-19, we further investigate the confirmed and death cases for the USA, China, UK, India and Brazil after their vaccination campaign starts. **Figure 7** presents the status of confirmed and death cases of five (5) most populated countries around the globe. Based on the findings, it is evident that the confirmed trend of USA was rising until January 20, 2021 and then gradually it decrease in passage of time. However, other countries have maintained a linear status in their confirmed cases but Brazil and India suddenly finds increasing number of COVID-19 cases after March 15, 2021. On the other hand, except Brazil all other countries maintained a descending trend in their death cases. Brazil started its vaccination on January 17, 2021 and still the country is fighting to minimize the death cases; on 18 march, 2021 it experienced a total death of almost 2,800 peoples. Overall, different vaccines have been proved effective against COVID-19 around the world and with the maintain of proper health regulation it may possible to reduce both the confirmed and death cases.

**Figure 7:**
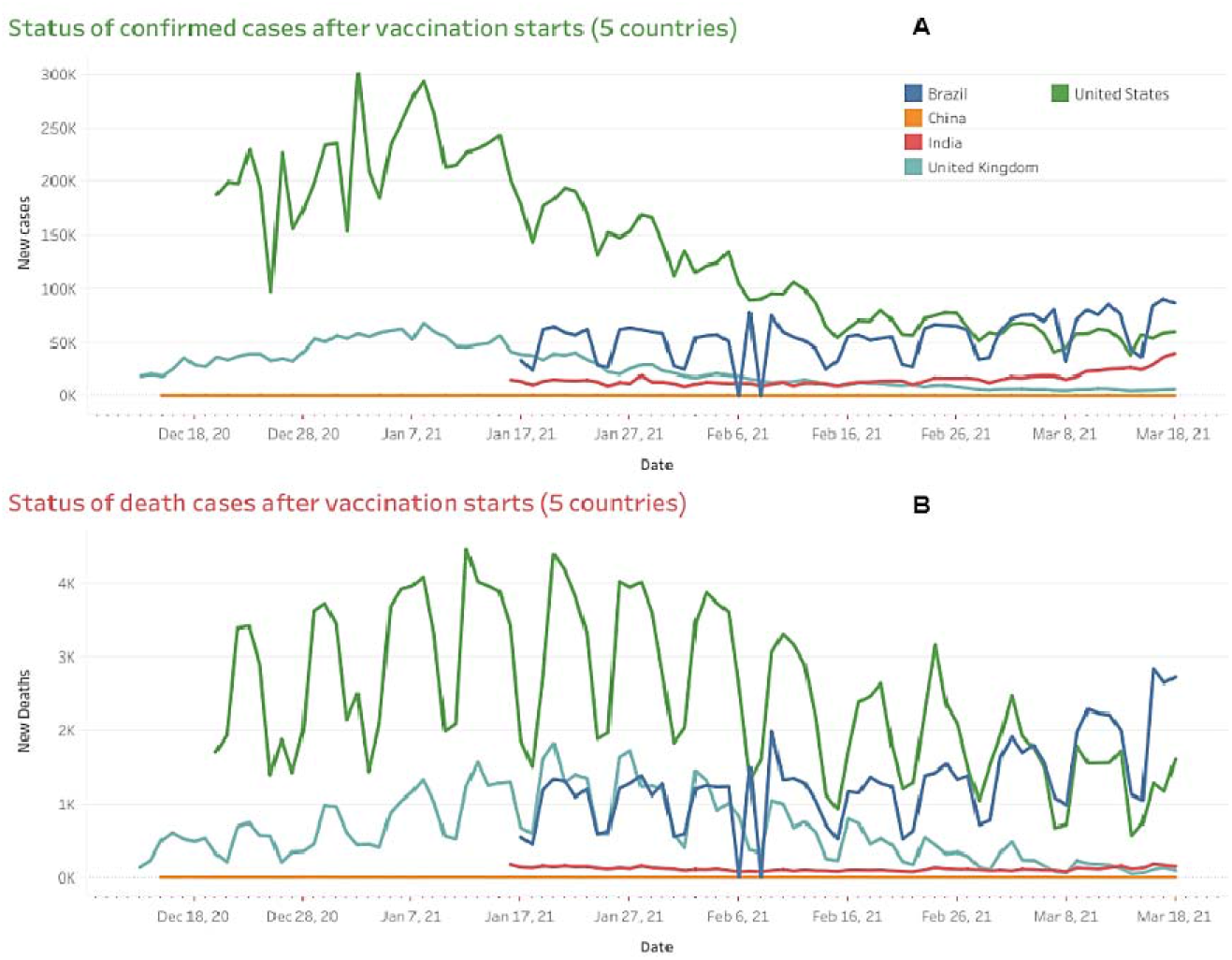
Confirmed and death cases analysis of five populated countries after its vaccination starts. 5(A) shows the confirmed cases of each countries until 18 March whereas 5(B) highlighting the death cases to analyze the impact of vaccination program.

This analysis also emphasized on the different vaccine production and their used in different context. Therefore, following **Table 5** is designed to provide an insight about each vaccine that is widely used and popular.

Based on the vaccination data available along with the support of Open Street Map and Map box countrywide map visualization is shown in **Figure 8**. The map is designed to provide insights of how different region and countries are ensuring vaccine for their populations. However, this map only highlights those regions that are currently offering vaccine. Surprisingly, few countries from the region of central Africa are still unable to ensure safe vaccine for their citizens. Until March 20, 2021, the United States and China accounted for about half of all vaccines used worldwide.

**Figure 8:**
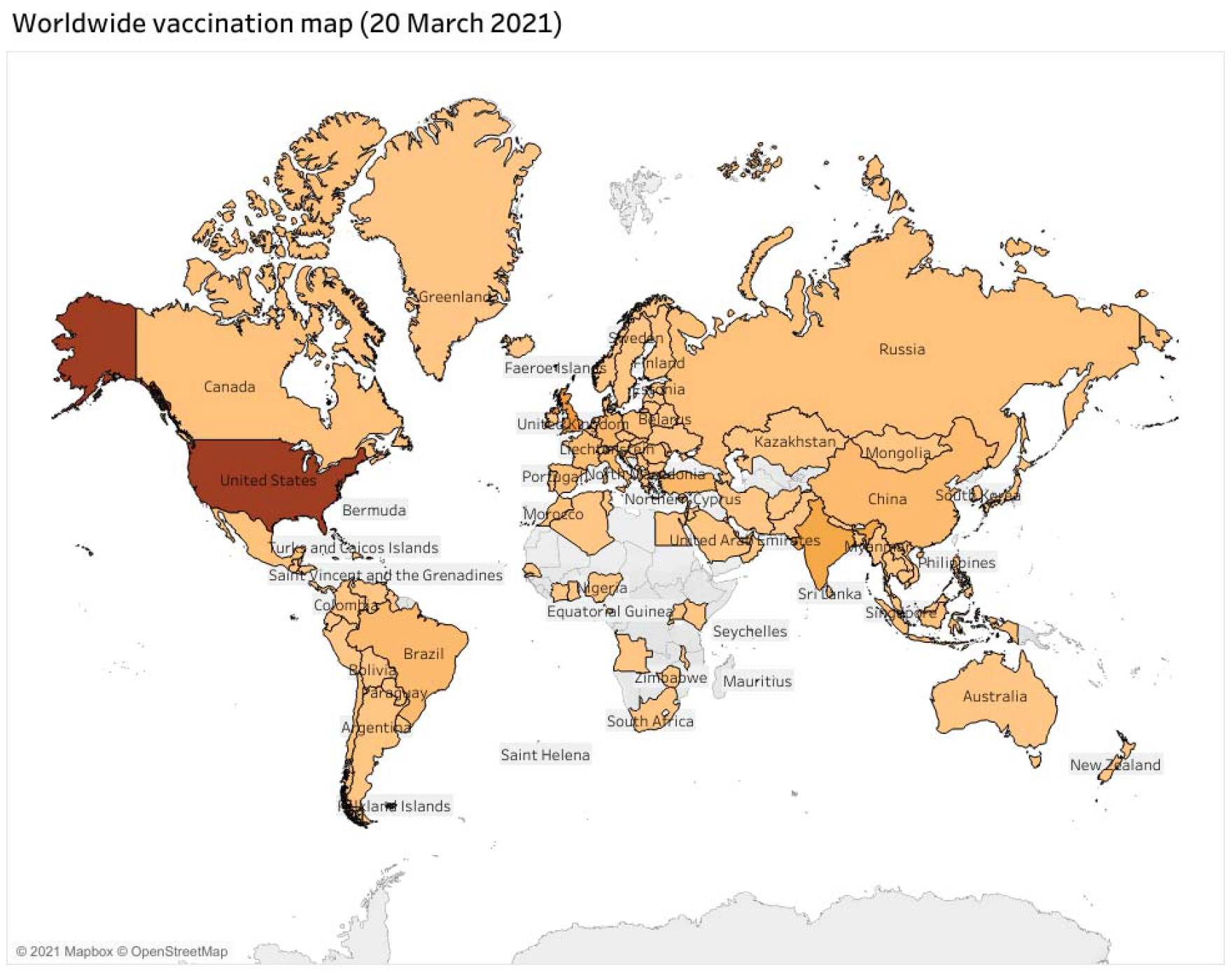
Worldwide vaccination map visualization based on highest number of doses offered by the country.

Until 21 March 2021, globally 150 countries are adopted 25 different vaccines for its population. Among them, some of the countries are using multiple vaccine for different categories of population according to their requirements. This research have unfolded the insights of vaccination program all over the world with data analysis and visualization. In addition, it tries to answers few questions regarding current status of different vaccine used in different region of the world. According to data analysis, Moderna, Pfizer/BioNTech, Oxford/AstraZeneca, Sinovac, Covaxin, and Covishield are the most popular vaccine used worldwide for mass vaccination, since all the vaccine has almost negligible side effects (known until 21 March 2021). This is quite pleasing that people from all the parts of the world are educating themselves and willingly taking the vaccines. In addition, scientist, public health experts, WHO, medical experts have claimed that these vaccines are effective against COVID-19 and that have already proven (21 March 2021). Apart from analyzing the findings, this study motivates a large number of people all over the world to take the vaccine and unite under one umbrella. If the vaccination rate continues to rise, all countries will be able to vaccinate their citizens by the end of the year.

## 5. Conclusion

The COVID-19 pandemic is still wreaking havoc on human lives around the world, but the COVID-19 vaccine offers a ray of hope for the future. The global response to the COVID-19 pandemic is likely to rely heavily on vaccine deployment. The present study revealed that globally there is a significant turnaround in people’s perception towards vaccination and thus different countries are ensuring mass vaccination for their citizens. The findings also suggests that, rate of vaccination among global population is increasing day by day and as such, unvaccinated peoples around the globe gets more motivated and encouraged to take vaccine in upcoming days. Although vaccines are still unavailable in some parts of the world for various reasons, we believe policymakers should take steps to ensure sufficient knowledge, positive attitudes, and perceptions of COVID-19 vaccinations in order to decrease vaccine hesitancy.

## Data Availability

The data that support the findings of this study are available from the corresponding author, upon reasonable request.

## Declaration

### Author Contribution

All authors conceptualized and designed the study. SKD and MR had the idea for and designed the study and had full access to all the data in the study and take the responsibility for the exploratory data analysis with their visualization. URS and AH and contributed to the writing of the article. MR contributed to the critical revision of the report. All the visualization and data presentation methods developed by SKD, MR, and AT. All authors contributed to data acquisition, data analysis, and reviewed and approved the final version.

### Ethical Approval

Not required

### Consent to participate

Not required

